# Body-worn inertial measurement units track knee flexion angles with similar accuracy to optical motion capture

**DOI:** 10.1101/2025.10.20.25338317

**Authors:** Ke Song, Josh R. Baxter

## Abstract

Knee motion is altered in overuse injuries and chronic diseases like osteoarthritis. Yet, relying on marker-based motion capture in the lab limits our understanding of how knee motion continuously impacts joint health in the real world. Markerless optical motion capture improves clinical suitability, but space and time constraints remain barriers toward real-world assessments. Inertial measurement unit (IMU) sensors have enabled continuous motion tracking outside the lab. In this study, we recorded thigh and shank-worn IMU data concurrently with marker-based and markerless optical motion capture on 10 healthy adults, who performed various daily living and exercise movements. We developed an IMU virtual alignment and data fusion paradigm to estimate knee flexion angle during each movement. We compared IMU-based estimate against marker-based and markerless motion capture using Pearson correlation (R_xy_) and root-mean-square difference (RMSD). IMU-estimated knee flexion angle strongly correlated with motion capture (R_xy_ ≥ 0.9). RMSDs were small for slower movements like walking, stairs, and squats (RMSD = 4.4° – 6.0°) while larger during faster movements like running and jumping (RMSD = 5.4° – 9.4°). Our findings show that wearable IMUs track knee flexion with similar accuracy to optical motion capture during daily living activities typical to older adults, highlighting their potential for monitoring real-world mobility in knees with chronic diseases. Conversely, it remains inconclusive whether IMUs accurately track dynamic knee motion relevant to athletic injuries. Future research should seek best practice for IMU wearing and mitigate practical pitfalls to secure high-fidelity data, for identifying clinically meaningful real-world biomarkers of knee mobility.

## 1. Introduction

Knee motion is critical for maintaining quality of life and long-term musculoskeletal health of the joint. It is altered both in chronic diseases like knee osteoarthritis (Baliunas et al., 2002; Astephen et al., 2008) and athletic overuse conditions like patellar tendinopathy (Rosen et al., 2015) and patellofemoral pain (Powers et al., 1999). Past studies have identified movement-related biomarkers for those conditions. For example, clinical gait analysis found that knee osteoarthritis reduces knee flexion during walking (Baliunas et al., 2002; Astephen et al., 2008). A recent study found that knee flexion increased in osteoarthritis patients who improve after joint replacement surgery, but not in patients who fail to improve (Amiri et al., 2023). However, most motion analyses rely on marker-based motion capture in the lab or clinic, while many only assess walking. Although markerless optical motion capture (Kanko et al, 2021; Uhlrich et al., 2023) has improved its suitability for clinical implementation, it often demands computation time and large data management (Edwards et al., 2025) while still being constrained by a fixed capture space. Isolated in-lab gait analysis does not capture the variability of real-world knee motion in daily living activities, nor the large variety of knee rehabilitation exercises (Song et al., 2023b). This limits our understanding of the chronic biomechanical impacts of knee motion, in context of both the natural history and rehabilitation outcomes for various knee conditions.

Inertial measurement unit (IMU) provides a new approach towards portable real-world motion capture (Seel et al., 2014; Hafer et al., 2023; Cereatti et al., 2024). Body-worn IMUs can be used to estimate body segment kinematics via accelerometer-gyroscope data fusion (Seel et al., 2014; Madgwick et al., 2011), which expands our ability to quantify joint movements outside of traditional laboratory and clinic settings (Hafer et al., 2023; Cereatti et al., 2024). While many studies have compared IMU-estimated knee flexion angle to marker-based (Kobsar et al., 2020), approaches and results have been highly variable. Few have compared both IMU and markerless optical systems concurrently to conventional marked-based motion capture; we are only aware of one such study, limited to 4 athletic activities (Edwards et al., 2025). No study has compared IMU and markerless optical motion capture directly to each other. Concurrent multi-system comparisons across slow (e.g., gait, daily living functions) and fast activities (e.g., running, sport exercises) will draw valuable insights into the population-specific pros and cons of IMU motion tracking compared to both field standards (marker-based) and alternative emerging technologies (markerless optical). These insights will help researchers, clinicians, and athletic trainers select the most suitable motion tracker based on their needs. Directly benchmarking IMU-based joint kinematics against both marker-based and markerless optical motion capture will also support translational studies that span multiple sites and combine in-lab with real-world assessments.

Our goal was to determine the concurrent validity between thigh and shank-worn IMUs and optical motion capture (marker-based and markerless) for estimating the knee flexion angle during various daily living and exercise movements. We hypothesized that knee flexion angle estimated from IMUs would be within the margin of difference between the marker-based and markerless optical systems, and kinematic waveforms would agree strongly across all 3 systems.

## 2. Methods

### 2.1. Study participant characteristics

We recruited 10 healthy young adults (5 male, 5 female, 21.9 ± 1.9 years old, height = 1.72 ± 0.09 m, mass = 71.4 ± 13.9 kg, body mass index = 23.8 ± 2.4 kg/m^2^) from our local community. We confirmed each study participant had no current or self-reported history of any lower limb or lower back injury that could interfere with daily living and athletic movements. All participants provided written informed consent before participating. This study was approved by the University of Pennsylvania Institutional Review Board (Protocol #850424) and performed in accordance with the Declaration of Helsinki and relevant human participant research guidelines.

### 2.2 Experimental procedure and knee flexion angle from optical motion capture

Study participants wore exercise clothing (running shorts and tank tops) and standardized running shoes that we provided. We attached a pair of research-grade IMUs with accelerometer and gyroscope (Opal V2R, APDM Inc., Portland, OR) to the right leg of each participant, one on the antero-lateral side of mid-thigh and the other on lateral mid-shank (**Figure 1A**). To minimize potential discomfort, we wrapped protective elastic bands around participant’s skin before using elastic straps to fasten each IMU securely over the protective bands. Both IMUs recorded at 100 Hz, using a ±16 g amplitude range for accelerometer and ±2000 °/s for gyroscope. The two IMUs were synchronized with each other via real-time Bluetooth wireless streaming to a data receiver (APDM Access Point). We then synchronized this data receiver with our optical motion capture systems through a vendor-provided synchronization module (APDM Sync Box v2).

**Figure 1.**
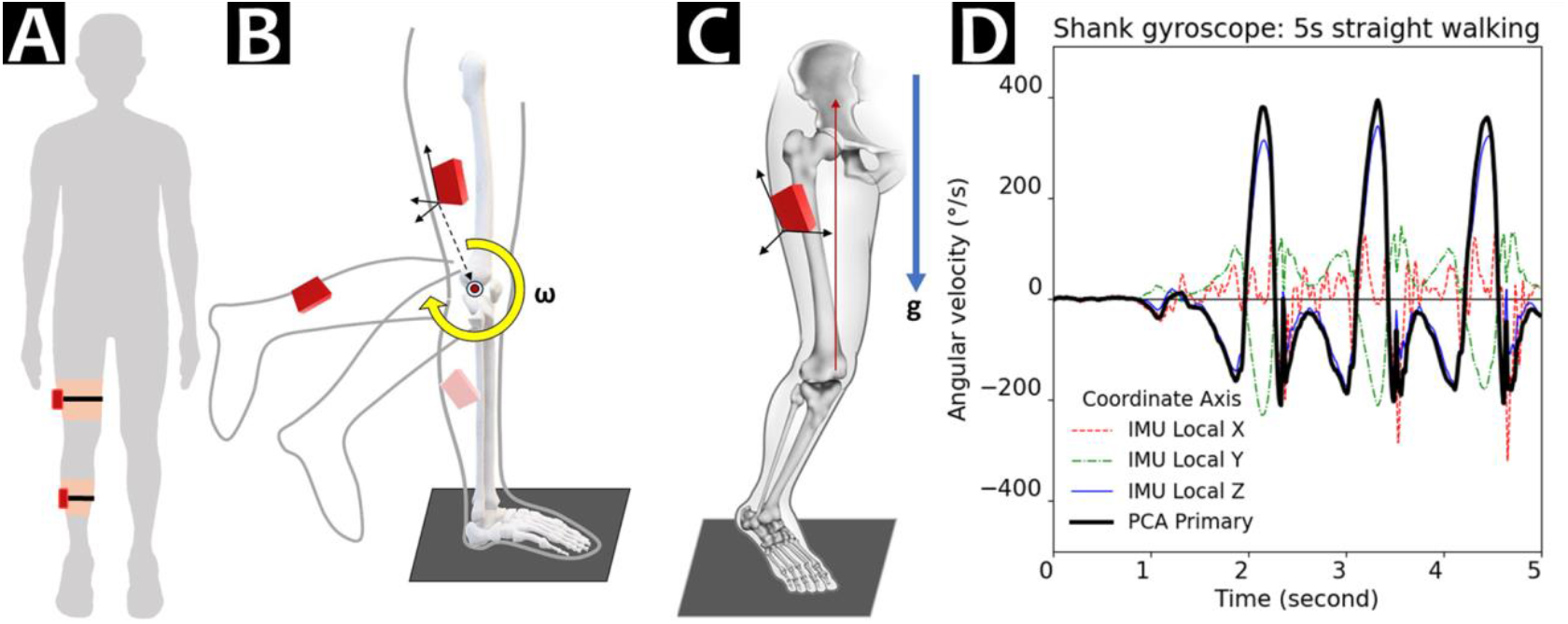
IMU physical placement and sensor-to-segment virtual realignment. **(A)** IMU locations on the right leg, one on lateral mid-thigh and the other on lateral mid-shank. **(B)** We used principal component analysis to identify the sagittal knee flexion axis, then **(C)** gravity at the static pose to identify the vertical axis. Diagrams (B) and (C) together depict our workflow for the sensor-to-segment alignment (reproduced from Cereatti et al., 2024, as permitted by a CC BY 4.0 license). **(D)** Example gyroscope data (angular velocity) during the 5-second straight walking bout used in principal component analysis (PCA). Although the local Z axis of this shank-worn IMU (blue solid line) roughly aligned with the knee flexion axis, principal component analysis-based transformation also integrated rotations left outside of Z axis (X: red dotted, Y: green dashed) to ensure the full knee flexion was captured by the virtually defined primary rotational axis (thick black solid line).

We first recorded a static pose trial with each participant standing in the anatomical position, with legs close together and knees fully extended. Each participant then performed 36 movements that are either activities of daily living or athletic exercises, in a preset order (**Appendix A**; Song et al., 2023b). We recorded at least 3 successful repetitions for each movement. We focused our main analysis on 10 representative movements (bolded texts in **Appendix A**), which are either the most essential tasks required for daily living or the most common rehabilitation exercises used by clinicians to treat knee pathologies. We repeated our analysis on the other 26 movements and reported them as secondary **Supplementary Data**.

We simultaneously collected optical motion capture data using both marker-based (1.3-megapixel Raptor-E ×10, 12-megapixel Raptor-12 ×2, Motion Analysis Corp., Rohnert Park, CA) and markerless cameras (2.1-megapixel Optitrack Prime Color, NaturalPoint Inc., Corvallis, OR) at 100 Hz. We previously published our optical motion capture setup, data collection and processing details, and comparison between concurrent markerless and marker-based joint angle estimates (Song et al., 2023a). Briefly, for marker-based, we secured 31 skin markers on the pelvis, thighs, shanks, and feet (on the shoes), and used a constrained kinematic model with 1 degree-of-freedom knee (flexion only; Slater et al., 2018) to track thigh and shank positions via marker trajectories. For markerless, we recorded synchronized videos and used a deep learning-based human motion tracker (Theia3D, Theia Markerless Inc., Kingston, ON, Canada) with 3 rotational degrees of freedom at the knee (Kanko et al., 2021a). The two optical motion capture systems and IMUs were all synchronized frame-by-frame via hardware triggering signals. We calculated knee flexion angle from both optical systems using inverse kinematics (Visual3D, C-Motion, Germantown, MD; Robertson et al., 2013), as previously described (Song et al., 2023a).

### 2.3. Knee flexion angle estimation using shank and thigh-worn IMUs

We developed a paradigm to estimate knee flexion angle from thigh and shank-worn IMUs using sensor-to-segment virtual alignment (**Figure 1B, 1C**) and accelerometer-gyroscope data fusion (**Appendix B**) (Hafer et al., 2020; Madgwick et al., 2011). First, we transformed accelerometer and gyroscope data from each IMU’s local frame (i.e., coordinate system) to an anatomically meaningful knee frame (Grood and Suntay, 1983). Sensor-to-segment coordinate transformation (Hafer et al. 2020) was required because the IMU local frames were orthogonal to the hardware surface and agnostic to knee anatomy, meaning none of the gyroscope signals readily represent knee flexion. To identify orientation of the knee flexion axis (X’) in the IMU local frame, we performed principal component analysis (**Figure 1B**) on each IMU’s gyroscope data during the first 5 seconds of the walking trial, when each participant walked along a straight path (**Figure 1D**). The first principal axis component where shank and thigh angular velocities were maximized during straight walking was our approximated knee flexion axis. We ensured this X’ virtual axis for both IMUs pointed laterally from the knee (i.e., same directional signs). Second, we identified the knee proximal-distal axis (Z) using average accelerometer data during the standing pose, with an assumption that both IMUs’ linear acceleration were aligned with gravity (**Figure 1C**). Third, we calculated the antero-posterior axis (Y) as the cross product of Z and X’ axes, then recalculated the final X axis to be the cross product of Y and Z axes for a fully orthogonal knee segment XYZ frame. We established IMU-to-segment frame rotational matrices independently for shank and for thigh, then transformed accelerometer and gyroscope data to their segment virtual frames, of both the X component approximates the projected knee flexion axis (Hafer et al., 2020) (**Figure 1D**).

We estimated IMU orientations using a commonly used open-source IMU sensor fusion algorithm (“Madgwick filter”; x-io Technologies Limited, Bristol, UK) to integrate gyroscope and accelerometer data (Madgwick et al., 2011; see **Appendix B** for fusion algorithm parameters and details). We chose this open-source algorithm over vendor-supplied proprietary fusion filter because it makes our paradigm applicable to other IMUs, especially low-cost devices that do not have a magnetometer. We converted fusion-estimated IMU orientations into Euler angles, where rotation around the first principal axis (X) represents the sagittal segment angle. We subtracted the sagittal shank and thigh angles for the knee flexion angle (Hafer et al., 2020). We calculated IMU-based knee flexion angle this way for each participant on each movement trial.

### 2.4. Data analysis and statistics

We computed the knee flexion angle during all movement repetitions from all 3 motion tracking systems. For cyclic gaits (walking and running), we identified gait cycles using ground reaction force and heel position; we were able to capture 13+ walking cycles and 4+ running cycles for all participants. For other motion trials, we manually defined start and stop events that represent the weight-bearing phase, as we previously detailed (Song et al., 2023b). We computed Pearson correlation coefficient (R_xy_) twice, one for IMU-based versus marker-based knee flexion estimate, and again separately for IMU versus the markerless optical estimate. R_xy_ quantifies the agreement of two knee flexion waveforms on each repetition. We also computed the root-mean-square difference (RMSD) separately for IMU versus marker-based and IMU versus markerless optical, which quantifies their mean magnitude differences. We previously reported concurrent validity between our marker-based and markerless optical systems from the same experiment (Song et al., 2023a); therefore, we did not include that comparison in our current work.

We averaged each R_xy_ and RMSD across trial repetitions within each participant. We then calculated the mean across 10 participants to determine the group-wise average agreement (R_xy_) and magnitude difference (RMSD) between IMU versus marker-based and IMU versus markerless optical estimates. According to the guidelines by Schober et al. (2018), we defined that R_xy_ ≥ 0.7 suggests a strong correlation between two systems, and R_xy_ ≥ 0.9 suggests a very strong correlation. To match recommendations in literature (Akbarshahi et al., 2010; Leardini et al., 2005) and our previous work (Song et al., 2023a), we defined that the joint angle magnitude difference is minimal if RMSD ≤ 5°.

## 3. Results

### 3.1. IMU tracking versus marker-based motion capture

Knee flexion angle estimated from thigh and shank-worn IMUs had excellent concurrent validity to marker-based estimates, especially during slower movements (**Figure 2**, IMU: red dashed, marker-based: black solid). Correlation of waveforms over a walking cycle was R_xy_ = 0.992, and their magnitude difference met our minimal criterion (RMSD = 4.4°). Likewise, during other slower movements like stair navigation (step up, step down) and high knee flexion tasks (squat, lunge), waveform correlations between IMU and marker-based were near perfect (R_xy_ ≥ 0.998), with magnitude differences just above our minimal criterion (RMSD: 5.4° – 6.0°). Differences were relatively larger for faster movements. Specifically, waveform correlation over a running gait cycle was R_xy_ = 0.990, and their magnitude difference exceeded our criterion of minimal (RMSD = 7.3°). Waveform correlations were excellent for vertical jump (R_xy_ = 0.997) and forward jump (R_xy_ = 0.992), while relatively less strong for repetitive jumps (R_xy_ = 0.977) and the run-and-cut maneuver (R_xy_ = 0.957). Magnitude difference for vertical jump (RMSD = 5.4°) was comparable to slower movements, but larger for forward jump, repetitive jumps, and run-and-cut (RMSD = 7.1°–7.7°). The most dynamic movement, run-and-cut, observed the largest magnitude difference (RMSD = 7.7°) and relatively the lowest waveform agreement (R_xy_ = 0.957), yet still met our a-priori criterion of very strong correlation.

**Figure 2.**
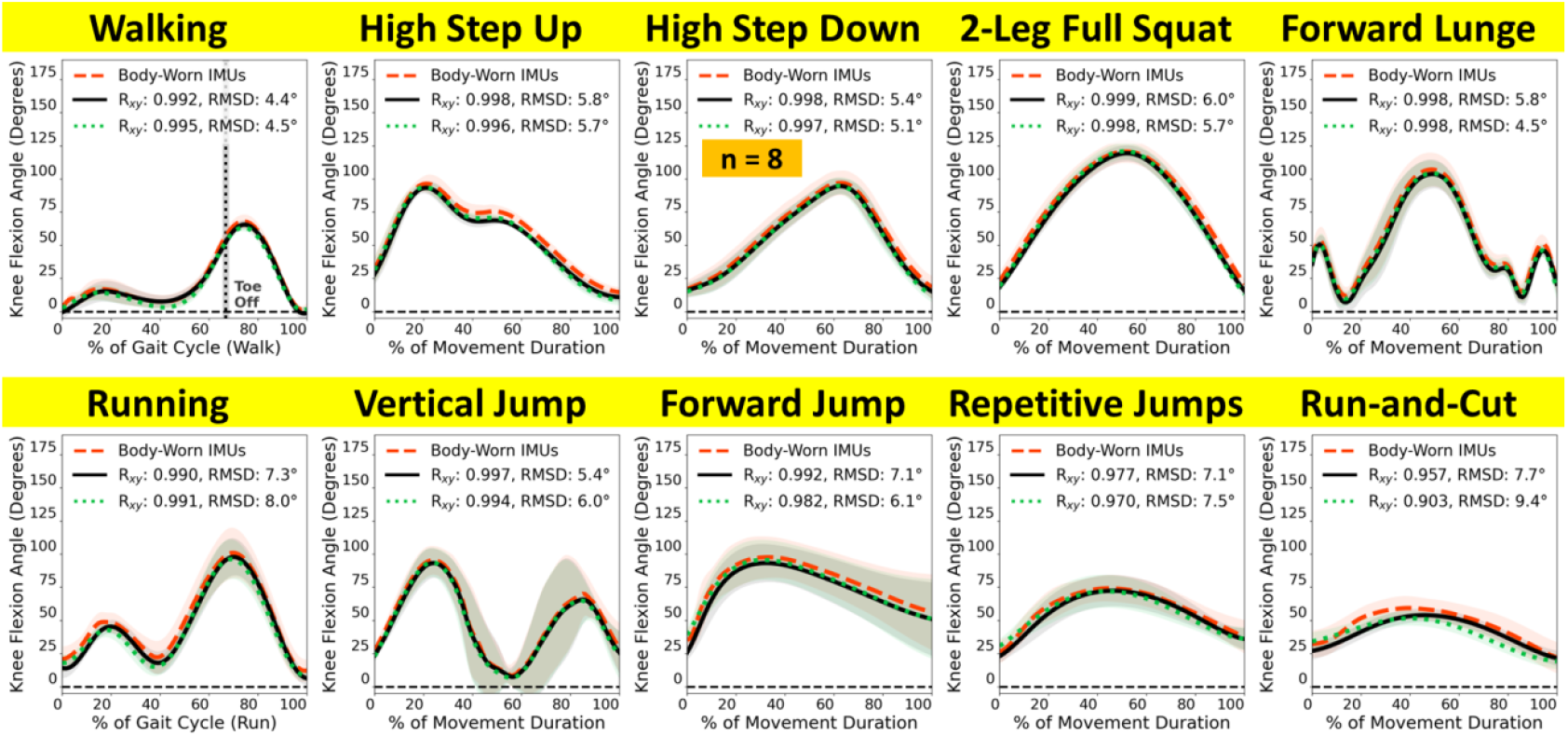
Knee flexion angle estimated from body-worn IMUs (red dashed) compared to marker-based (black solid) and markerless optical motion capture (green dotted) during 10 representative movements. Waveforms = group mean (line) ± 1 standard deviation (shade). **(Top)** IMUs had excellent agreements to both optical systems during gait, stair navigation, and high knee flexion movements. **(Bottom)** Differences between IMUs and the optical systems were larger for faster athletic movements like running and jumping. Walking and running were analyzed over a gait cycle (between heel strikes). Start and end definition for the other movements were detailed in our prior work (Song et al., 2023b). IMU data for high step down was lost for 2 participants due to technical errors.

### 3.2. IMU tracking versus markerless optical motion capture

The concurrent validity between IMU and our markerless optical system was generally the same as IMU versus marker-based: very strong waveform agreement and small magnitude differences, especially for slower movements (**Figure 2**, IMU: red dashed, markerless optical: green dotted). For a walking cycle, waveform correlation was R_xy_ = 0.995, and magnitude difference was RMSD = 4.5°. For other slower movements (stair navigation and high knee flexion), waveform agreements were again near-perfect (R_xy_ ≥ 0.996), and magnitude differences were likewise just above the minimal threshold (RMSD = 5.1°–5.7°) except for forward lunge, which met the minimal criterion (RMSD = 4.5°). For a running gait cycle, waveform agreement was almost the same as versus marker-based (R_xy_ = 0.991) while magnitude difference was marginally larger (RMSD = 8.0°). Waveform agreements were excellent for vertical jump (R_xy_ = 0.994) but slightly lower than the agreements with marker-based for other fast movements (R_xy_ = 0.903 – 0.982); magnitude differences varied in the above-minimal range (RMSD = 6.0° – 9.4°). Run-and-cut again observed the largest RMSD (9.4°) and the lowest waveform agreement (0.903), both relatively worse than the comparability between IMU and marker-based. Overall, across the 10 movements, walking consistently showed the best match between IMU and the two optical systems, while run-and-cut matched consistently the worst, followed by running.

## 4. Discussion

Our goal was to determine the concurrent validity between thigh and shank-worn IMUs and optical motion capture (marker-based and markerless) for estimating the knee flexion angle during various daily living and exercise movements. Results supported our hypotheses that knee flexion angle estimated using IMUs is within the margin of difference between marker-based and markerless optical motion capture, and that kinematic waveforms agreed strongly across all 3 systems. Magnitude differences were small during walking (<5°) and other slow movements (5– 6°) but larger for faster sports-related movements (5.4–9.4°). Excellent agreements during slower movements like gait support IMUs as a suitable tool for tracking daily living knee mobility in chronic disease populations like osteoarthritis. Conversely, it is inconclusive whether IMUs can accurately track dynamic knee motion relevant to athletic injuries. Our 3-way concurrent comparison between conventional marker-based motion capture, low-burden markerless motion capture, and wearable IMU motion tracking across slow and fast movements provide a first-of-kind benchmark for researchers, clinicians, and athletic trainers to evaluate the pros and cons of each platform and choose the best system for knee motion analysis based on their unique needs.

Our main finding was that IMUs track knee flexion during a full walking gait cycle with excellent waveform agreement (R_xy_ > 0.99) and minimal magnitude difference (RMSD < 5°) to both marker-based and markerless optical motion capture (**Figure 2**, top left). Our prior work reported from this same dataset that knee flexion differed by 3.9° between marker-based and markerless optical motion capture during walking stance (Song et al., 2023a). Our current results show that IMU-estimated knee flexion is within this difference margin between our two optical systems. Considered together with our prior work, markerless videos and leg-worn IMUs are both suitable for quantifying knee flexion and offer practical benefits over conventional marker-based systems. Markerless optical motion capture is a good choice for in-clinic motion analysis where laboratory space is available, due to its low burdens on patients and excellent inter-session reliability (Kanko et al., 2021b; Outerleys et al., 2024). IMUs are a good choice for clinical studies where laboratory space is limited, or when real-world continuous monitoring is essential.

We likewise found strong agreements between IMUs and optical motion capture during common daily living activities like stair navigation (step up and down) and high knee flexion movements (squat and lunge) (**Figure 2**, top row). These essential everyday functions place high mechanical demands on the knee joint, which patients with symptomatic knee osteoarthritis often struggle to perform. To understand how much total mechanical exposure is imposed on the diseased knee due to these movements, it is important to consider motion patterns together with the frequency of such motions in daily living. Together with walking, we demonstrate that high-fidelity IMU knee flexion tracking captures most daily activities performed in populations with average or limited knee mobility. We advocate for researchers to scale up IMU-based motion analysis studies, generate longer-duration data, and validate newly developed real-world motion parameters both technically (versus gold-standard motion analyses) and clinically (versus patient outcomes) to confirm the clinical values of real-world wearable motion sensing.

IMU-based knee flexion measured during faster movements strongly correlated with the optical systems (R_xy_ > 0.9), but RMSDs exceeded our 5° minimal difference criterion. Because soft-tissue artifacts (Leardini et al., 2005) can induce up to 8.3° error in marker-estimated knee flexion when compared to ground-truth biplane fluoroscopy (Akbarshahi et al., 2010), RMSD < 8° should be considered within the range of uncertainty when using marker-based motion capture as reference. Our IMU-based estimates were within this ∼8° uncertainty range versus marker-based across all 10 representative movements (**Figure 2**; RMSD ≤ 7.7°). We are not aware of any concurrent validation between leg-worn IMUs and biplane fluoroscopy, and it is beyond our scope to speculate whether IMU or marker-based knee flexion angle is closer to the ground truth for fast movements. The decision to trust IMUs in tracking knee flexion during those movements should thus be left to the confidence investigators have in optical motion capture fidelity.

Our results compare favorably to other IMU-based studies. Previous IMU studies that quantified knee flexion during walking and running reported RMSDs < 4° (Seel et al., 2014; Cooper et al., 2009; McGrath and Sterling, 2022; Rhudy et al., 2024). Our results matched those findings on walking (RMSD = 4.4°), although we saw a larger difference on running (7.3°). Studies on other knee exercises were highly variable on the reported inter-system differences. For example, RMSDs between IMU and marker-based during run-and-cut ranged from 1.1° (Fan et al. 2021) to 12.1° (Edwards et al. 2025); our finding (7.7°) falls within this range. Of note, although IMU and markerless optical tracking both gained traction in recent research, to our knowledge no study has directly compared concurrent knee flexion estimates between these two systems. In our results, the IMU-versus-markerless differences were almost the same as IMU-versus-marker-based (**Figure 2**). While each system estimates knee flexion comparably, especially for slower movements, they possess distinct practical strengths and limitations that should be evaluated when selecting the most suitable system for research or clinical purposes (Edwards et al., 2025).

Our findings should be interpreted with several limitations. Our sample size was limited to 10 healthy young adults, as we focused on establishing a first-of-its-kind 3-way concurrent validation dataset between IMU-based and optical motion analyses across a large variety of movements. We prioritized knee flexion, because frontal and transverse-plane knee kinematics estimated by marker-based motion capture are highly susceptible to skin artifacts (Akbarshahi et al., 2010) and are not reliable references for cross-validation. We defined full knee extension – required for defining virtual segment frames (**Figure 1C**) – using a standing pose trial. Standing with a straight knee may be difficult for patients with osteoarthritis due to painful limited motion. New experimental procedures to establish segment coordinate frames are needed, and the updated IMU algorithms should be revalidated in patients with symptomatic knee osteoarthritis. Our current experiments were performed in a lab, where secure sensor attachment was confirmed and data duration was short. This was necessary for validating against concurrent lab-based optical motion capture. Technical barriers remain for translating the good comparability we saw to reliable real-world continuous motion tracking, where repositioned sensors would require event detection and virtual recalibration (Cereatti et al., 2024; Cain and Morrow, 2025). Multi-sensor re-synchronization is also needed for long-duration tracking, where sensor internal clocks are impacted by quartz oscillator errors and compounded by ambient temperature (Walls and Gagnepain, 1992). Lower-cost IMUs for multi-day usages are commercially available but often lack sensor synchronization features, thus time-wise drift will emerge over week-long recordings that complicate accurate kinematic estimation (Cereatti et al., 2024; Cain and Morrow, 2025).

In conclusion, we developed a new thigh and shank-worn IMU paradigm that tracks knee flexion angles with similar accuracy to concurrent marker-based and markerless optical motion capture. Very strong agreements between IMUs and optical systems during essential daily living tasks like gait, stair navigation, and high knee flexion movements suggest that IMUs are suitable for monitoring the real-world mobility in knees with chronic diseases. Conversely, the accuracy of IMUs to track real-world dynamic knee motion relevant to athletic injuries is inconclusive and warrants further research. IMUs present unique challenges like sensor repositioning, syncing, and wearer comfort. Future work should develop strategies to mitigate these challenges and improve the feasibility, reliability, and validity of body-worn IMU continuous tracking of real-world knee motion. IMUs present an exciting new platform for clinicians to monitor real-world knee function in both physically active and impaired populations, for deeper insights into the cumulative impacts of knee motion on joint overuse injuries and chronic degenerative diseases.

## Data Availability

The original and processed IMU data, optical motion capture data, and biomechanical models that support the findings of this study are available from the corresponding author (K.S.) or the senior author (J.R.B.) upon reasonable request.

## CRediT authorship contribution statement

**Ke Song:** Conceptualization, Data curation, Formal analysis, Investigation, Methodology, Software, Validation, Visualization, Writing – original draft, Writing – review & editing.

**Josh R. Baxter:** Conceptualization, Funding acquisition, Methodology, Project administration, Resources, Supervision, Writing – review & editing.

## Acknowledgments

This work was funded by the National Institutes of Health NIAMS R01AR078898 and United States Department of Veterans Affairs VA I50RX004845. The funders had no role in the study design, collection, analysis and interpretation of data, writing of the report and decision to submit the article for publication. We thank Dr. Karin Grävare Silbernagel and Dr. Rodrigo Scattone Silva for experimental design of the original larger study, and Todd Hullfish, Madison Woods, Audrey Lehneis, and Liliann Zou for assistance with data collection and processing. We also thank Dr. Jocelyn Hafer for valuable guidance on IMU data processing and verification.

## Supplementary data

Supplementary data for this study can be found in the online version of this article.

## Appendix A: The 36 movement trials, listed in the order of experimental data collection

| Module #1<br>Exercise 1–11 | Module #2<br>Exercise 12–21 | Module #3<br>Exercise 22–29 | Module #4<br>Exercise 30–36 |
| --- | --- | --- | --- |
| Heel raises | **Running** | 1-leg squat (60-degree) | 1-leg drop landing |
| **Walking** | 2-leg squat (60-degree) | 1-leg squat (full depth) | 1-leg drop vertical hop |
| Low step up (10-cm) | **2-leg squat (full depth)**<br>**“2-Leg Full Squat”** | Bulgarian squat | 1-leg repeat forward hops |
| Low step down (10-cm) | 1-leg decline squat | 1-leg countermovement hop | 1-leg fast forward hops |
| **High step up (20-cm)** | Sumo squat | **2-leg repeat forward jumps**<br>**“Repetitive Jumps”** | 1-leg repeat lateral hops |
| **High step down (20-cm)** | 1-second Spanish squat | 2-leg fast forward jumps | 1-leg fast lateral hops |
| **Lunge**<br>**“Forward Lunge”** | **Run-and-cut** | 2-leg repeat lateral jumps | Alternating split jumps |
| **2-leg countermovement jump**<br>**“Vertical Jump”** | 1-leg maximal forward hop | 2-leg fast lateral jumps |  |
| 2-leg drop landing | Run-and-stop |  |  |
| 2-leg drop vertical jump | Sports movement jump |  |  |
| **2-leg maximal forward jump**<br>**“Forward Jump”** |  |  |  |

**Bolded texts** annotate the 10 movements reported as main findings in Results. The other 26 movements are reported as **Supplementary Data**. Detailed trial definitions can be found in Song et al. (2023b).

## Appendix B: Accelerometer-gyroscope fusion algorithm

The working mechanism of accelerometer-gyroscope fusion is well documented (Madgwick, 2011; Seel et al., 2014). Briefly, we integrated angular velocity from gyroscope to estimate orientation (angle), and used linear acceleration from accelerometer to correct for sensor tilt relative to gravity and the drift accumulated from gyroscope noise. We set the gain of the Madgwick filter at *β* = 0.5. We noticed during pilot testing that our filter took up to 5 seconds to converge from an arbitrary initial orientation guess. As many of our trials were brief, to ensure correct estimation for the whole trials, we prepended 5 seconds of virtual IMU data that “froze” at linear acceleration of the start of each trial, with zero angular velocity. This “freeze” allowed our algorithm to coverage to its true starting orientation in time. Output of the fusion filter was the orientation of each IMU relative to global inertial frame expressed as quaternions.

