## Supplementary Data for "Body-worn inertial measurement units track knee flexion angles with similar accuracy to optical motion capture"

**Supplementary Data.** Knee flexion angle estimated from body-worn IMUs compared to marker-based and markerless optical systems across a total of 36 movements.

The secondary data presented herein are extended from the same biomechanical analysis we performed on the 10 movements presented in the main article. Each subject performed all 36 movements during the experiment of a larger study, in the order identical to the Supplementary Figures presented below (i.e., heel raises, walking, etc.) **Titles highlighted in teal color** annotate the 10 representative movements we included in the main article. Waveforms = group mean (line) = ± 1 standard deviation (shade) for **body-worn IMUs (red dashed)**, **marker-based (black solid)**, and **markerless optical (green dotted)** systems. Start and end definition for the other movements were detailed in our prior work (Song et al., 2023b). IMU data for some participants were lost for low step down (1 lost), high step down (2 lost), 2-leg drop vertical jump (1 lost), and 2-leg fast-speed lateral jumps (1 lost) due to technical errors during the experiment (labeled “n = #”).

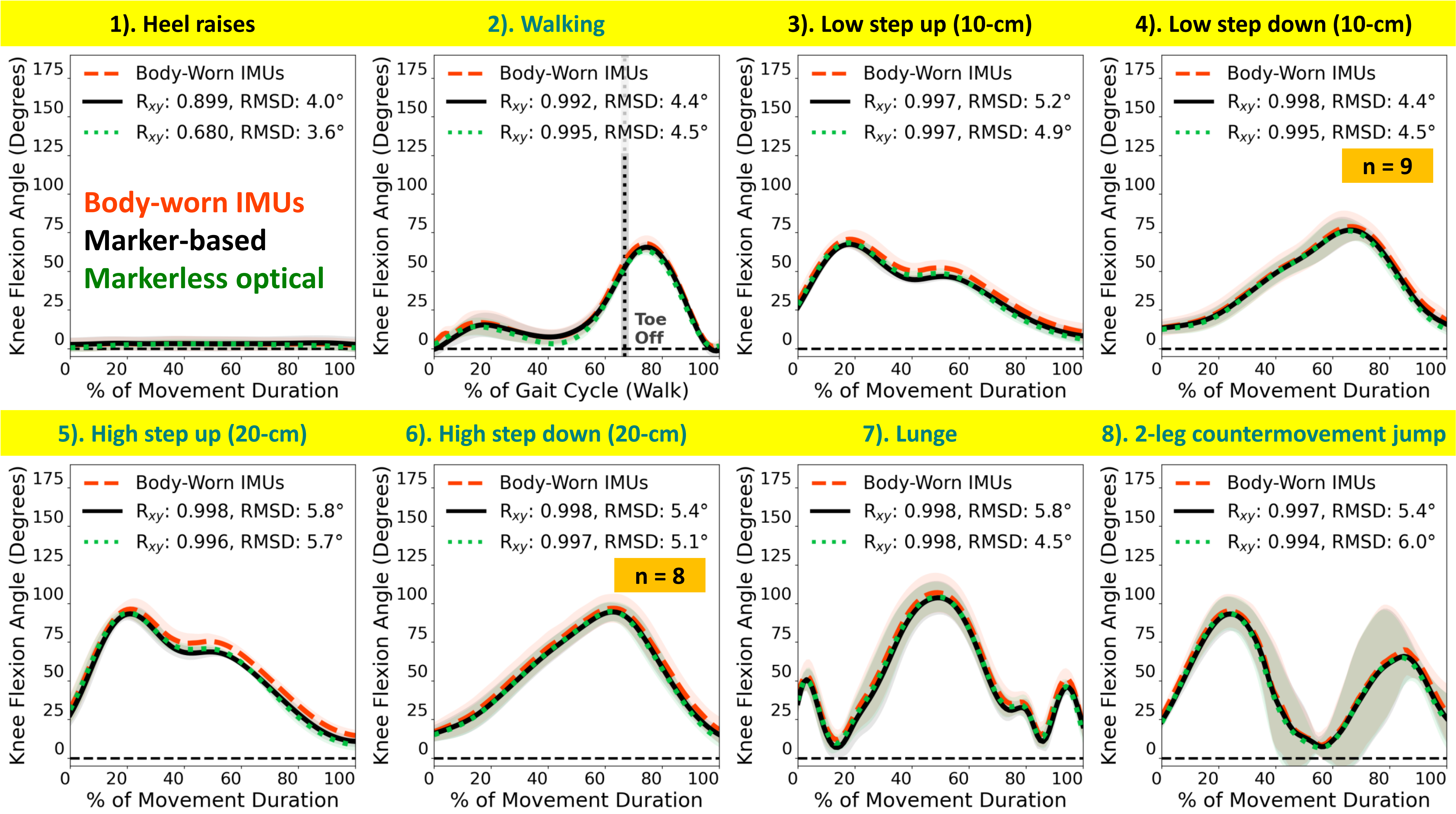
1). Heel raises
2). Walking
3). Low step up (10-cm)
4). Low step down (10-cm)
5). High step up (20-cm)
6). High step down (20-cm)
7). Lunge
8). 2-leg countermovement jump
Body-worn IMUs
Marker-based
Markerless optical
Body-Worn IMUs
R$_{xy}$: 0.899, RMSD: 4.0°
R$_{xy}$: 0.680, RMSD: 3.6°
R$_{xy}$: 0.992, RMSD: 4.4°
R$_{xy}$: 0.995, RMSD: 4.5°
Toe Off
R$_{xy}$: 0.997, RMSD: 5.2°
R$_{xy}$: 0.997, RMSD: 4.9°
R$_{xy}$: 0.998, RMSD: 4.4°
R$_{xy}$: 0.995, RMSD: 4.5°
n = 9
R$_{xy}$: 0.998, RMSD: 5.8°
R$_{xy}$: 0.996, RMSD: 5.7°
R$_{xy}$: 0.998, RMSD: 5.4°
R$_{xy}$: 0.997, RMSD: 5.1°
n = 8
R$_{xy}$: 0.998, RMSD: 5.8°
R$_{xy}$: 0.998, RMSD: 4.5°
R$_{xy}$: 0.997, RMSD: 5.4°
R$_{xy}$: 0.994, RMSD: 6.0°
Knee Flexion Angle (Degrees)
% of Movement Duration
% of Gait Cycle (Walk)

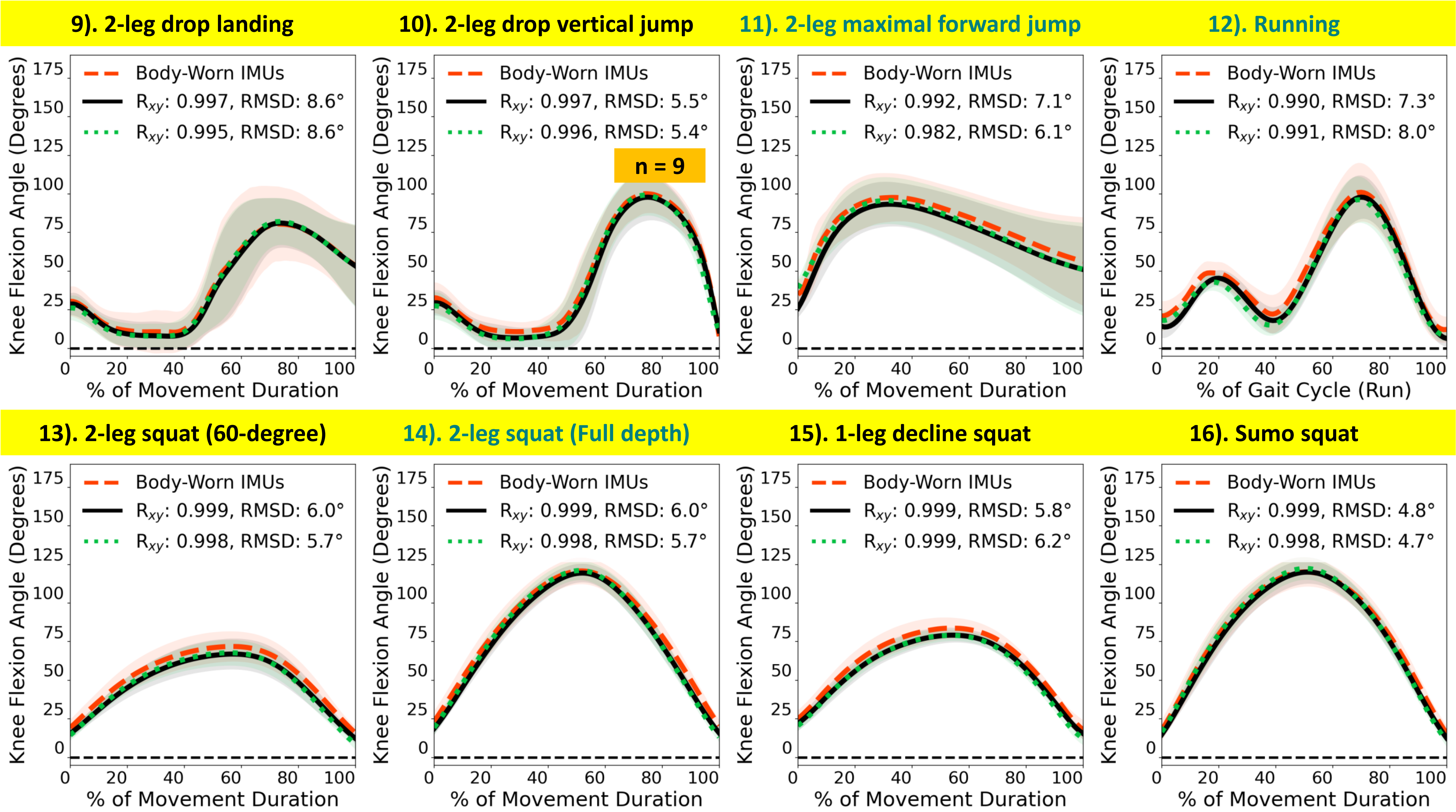
9). 2-leg drop landing
Body-Worn IMUs
$R_{xy}$: 0.997, RMSD: 8.6°
$R_{xy}$: 0.995, RMSD: 8.6°
10). 2-leg drop vertical jump
Body-Worn IMUs
$R_{xy}$: 0.997, RMSD: 5.5°
$R_{xy}$: 0.996, RMSD: 5.4°
n = 9
11). 2-leg maximal forward jump
Body-Worn IMUs
$R_{xy}$: 0.992, RMSD: 7.1°
$R_{xy}$: 0.982, RMSD: 6.1°
12). Running
Body-Worn IMUs
$R_{xy}$: 0.990, RMSD: 7.3°
$R_{xy}$: 0.991, RMSD: 8.0°
% of Gait Cycle (Run)
13). 2-leg squat (60-degree)
Body-Worn IMUs
$R_{xy}$: 0.999, RMSD: 6.0°
$R_{xy}$: 0.998, RMSD: 5.7°
14). 2-leg squat (Full depth)
Body-Worn IMUs
$R_{xy}$: 0.999, RMSD: 6.0°
$R_{xy}$: 0.998, RMSD: 5.7°
15). 1-leg decline squat
Body-Worn IMUs
$R_{xy}$: 0.999, RMSD: 5.8°
$R_{xy}$: 0.999, RMSD: 6.2°
16). Sumo squat
Body-Worn IMUs
$R_{xy}$: 0.999, RMSD: 4.8°
$R_{xy}$: 0.998, RMSD: 4.7°
Knee Flexion Angle (Degrees)
% of Movement Duration
0
25
50
75
100
125
150
175
0
20
40
60
80
100

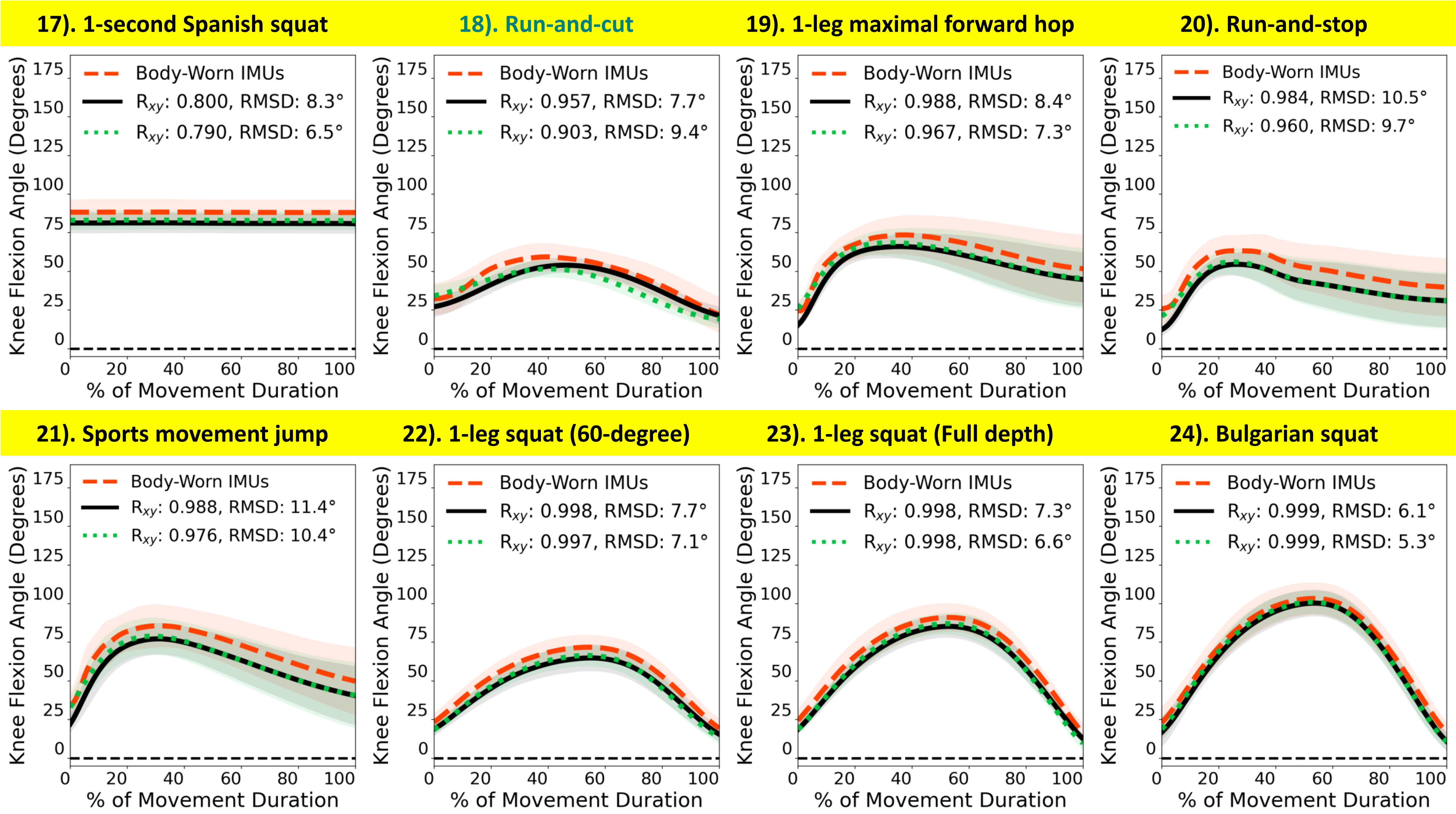
17). 1-second Spanish squat
Body-Worn IMUs
$R_{xy}$: 0.800, RMSD: 8.3°
$R_{xy}$: 0.790, RMSD: 6.5°
18). Run-and-cut
Body-Worn IMUs
$R_{xy}$: 0.957, RMSD: 7.7°
$R_{xy}$: 0.903, RMSD: 9.4°
19). 1-leg maximal forward hop
Body-Worn IMUs
$R_{xy}$: 0.988, RMSD: 8.4°
$R_{xy}$: 0.967, RMSD: 7.3°
20). Run-and-stop
Body-Worn IMUs
$R_{xy}$: 0.984, RMSD: 10.5°
$R_{xy}$: 0.960, RMSD: 9.7°
21). Sports movement jump
Body-Worn IMUs
$R_{xy}$: 0.988, RMSD: 11.4°
$R_{xy}$: 0.976, RMSD: 10.4°
22). 1-leg squat (60-degree)
Body-Worn IMUs
$R_{xy}$: 0.998, RMSD: 7.7°
$R_{xy}$: 0.997, RMSD: 7.1°
23). 1-leg squat (Full depth)
Body-Worn IMUs
$R_{xy}$: 0.998, RMSD: 7.3°
$R_{xy}$: 0.998, RMSD: 6.6°
24). Bulgarian squat
Body-Worn IMUs
$R_{xy}$: 0.999, RMSD: 6.1°
$R_{xy}$: 0.999, RMSD: 5.3°
Knee Flexion Angle (Degrees)
% of Movement Duration

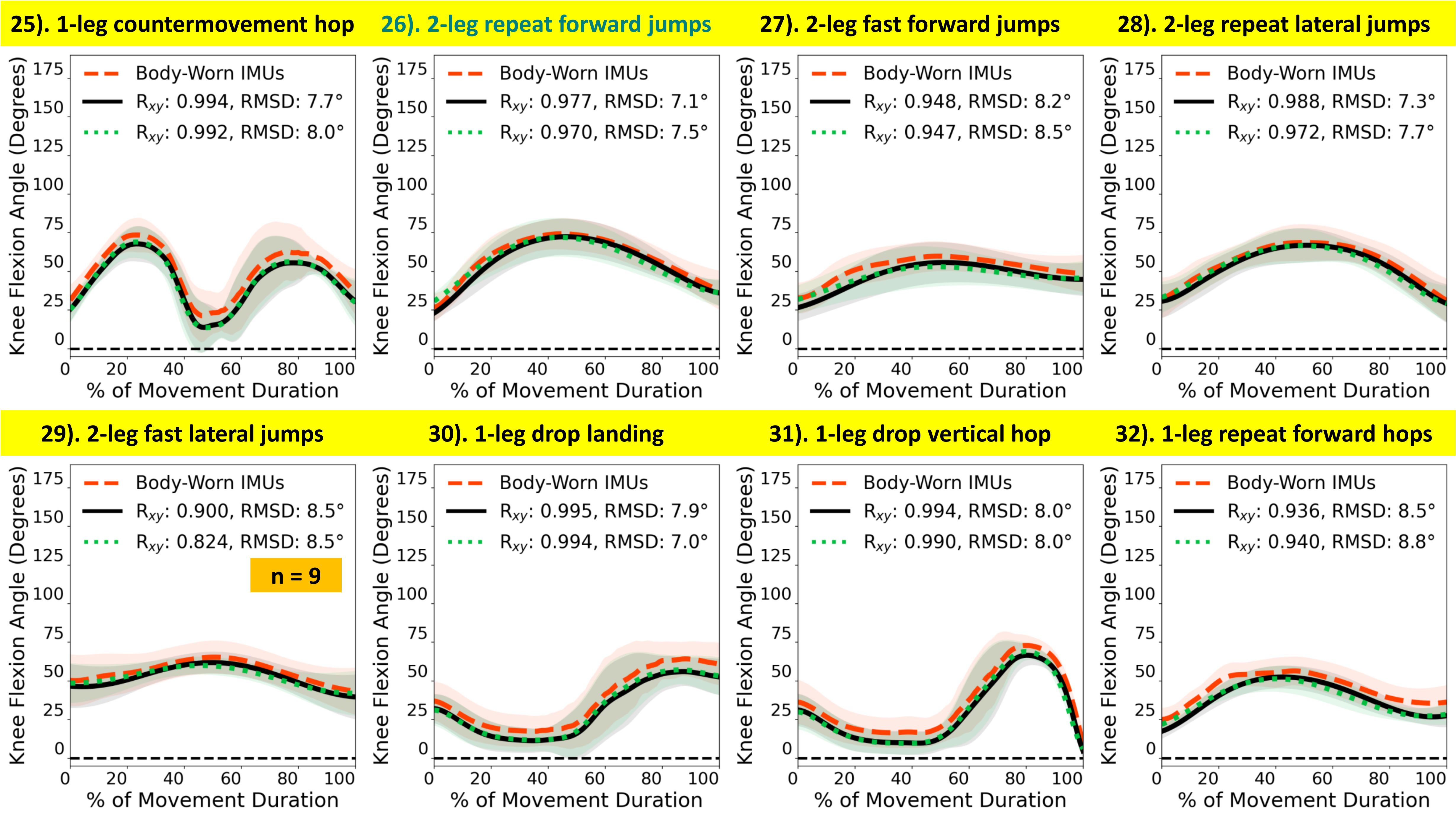
25). 1-leg countermovement hop
Body-Worn IMUs
Rxy: 0.994, RMSD: 7.7°
Rxy: 0.992, RMSD: 8.0°
26). 2-leg repeat forward jumps
Body-Worn IMUs
Rxy: 0.977, RMSD: 7.1°
Rxy: 0.970, RMSD: 7.5°
27). 2-leg fast forward jumps
Body-Worn IMUs
Rxy: 0.948, RMSD: 8.2°
Rxy: 0.947, RMSD: 8.5°
28). 2-leg repeat lateral jumps
Body-Worn IMUs
Rxy: 0.988, RMSD: 7.3°
Rxy: 0.972, RMSD: 7.7°
29). 2-leg fast lateral jumps
Body-Worn IMUs
Rxy: 0.900, RMSD: 8.5°
Rxy: 0.824, RMSD: 8.5°
n = 9
30). 1-leg drop landing
Body-Worn IMUs
Rxy: 0.995, RMSD: 7.9°
Rxy: 0.994, RMSD: 7.0°
31). 1-leg drop vertical hop
Body-Worn IMUs
Rxy: 0.994, RMSD: 8.0°
Rxy: 0.990, RMSD: 8.0°
32). 1-leg repeat forward hops
Body-Worn IMUs
Rxy: 0.936, RMSD: 8.5°
Rxy: 0.940, RMSD: 8.8°
Knee Flexion Angle (Degrees)
% of Movement Duration

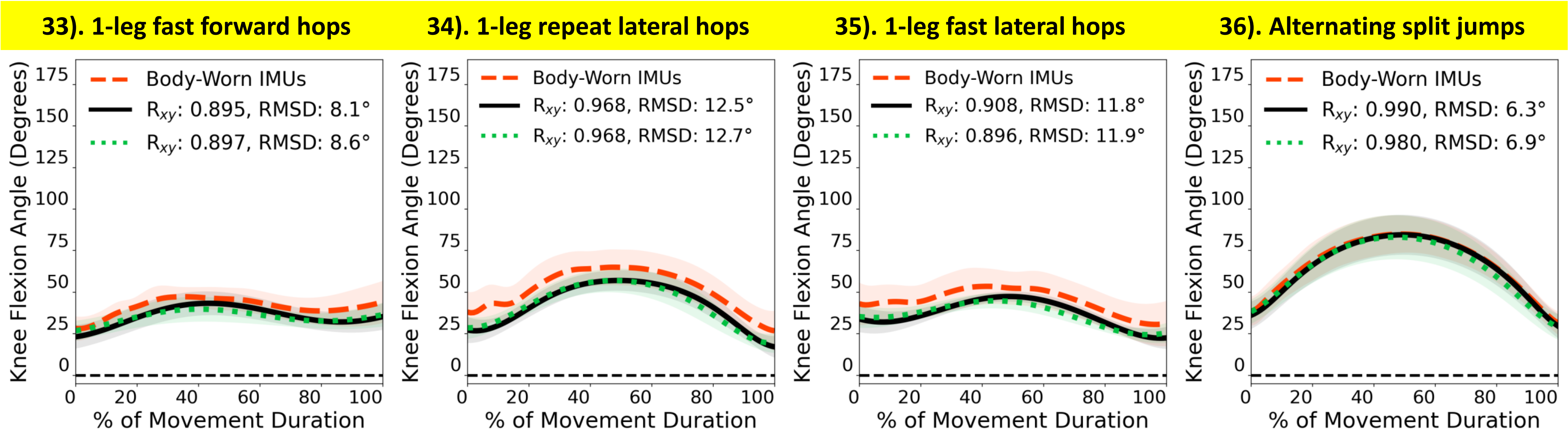
33). 1-leg fast forward hops
34). 1-leg repeat lateral hops
35). 1-leg fast lateral hops
36). Alternating split jumps
Body-Worn IMUs
Rxy: 0.895, RMSD: 8.1°
Rxy: 0.897, RMSD: 8.6°
Body-Worn IMUs
Rxy: 0.968, RMSD: 12.5°
Rxy: 0.968, RMSD: 12.7°
Body-Worn IMUs
Rxy: 0.908, RMSD: 11.8°
Rxy: 0.896, RMSD: 11.9°
Body-Worn IMUs
Rxy: 0.990, RMSD: 6.3°
Rxy: 0.980, RMSD: 6.9°
Knee Flexion Angle (Degrees)
% of Movement Duration
